# COVID-19 Diagnostic Testing For All - Using Non-Dilutive Saliva Sample Collection, Stabilization and Ambient Transport Devices

**DOI:** 10.1101/2021.01.20.20243782

**Authors:** Yuan Carrington, Justin Orlino, Alejandro Romero, Jessica Gustin, Mahssa Rezaei, Elizabeth Greene, Summer Rose, Ramani A. Aiyer, Shanavaz Nasarabadi

## Abstract

COVID-19 testing is not accessible for millions during this pandemic despite our best efforts. Without greatly expanded testing of asymptomatic individuals, contact tracing and subsequent isolation of spreaders remains as a means for control. In an effort to increase RT-PCR assay testing for the presence of the novel beta-coronavirus SARS-CoV-2 as well as improve sample collection safety, GenTegra LLC has introduced two products for saliva collection and viral RNA stabilization: GTR-STM™ (GenTegra Saliva Transport Medium) and GTR-STMdk™ (GenTegra Saliva Transport Medium Direct to PCR). Both products contain a proprietary formulation based on GenTegra’s novel “Active Chemical Protection™” (ACP) technology that gives non-dilutive, error-free saliva sample collection using RNA stabilization chemicals already dried in the collection tube.

GTR-STM can be used for safer saliva-based sample collection at home (or at a test site). Following saliva collection, the sample-containing GTR-STM can be kept at ambient temperature during shipment to an authorized CLIA lab for analysis. SARS-CoV-2 viral RNA in GTR-STM is stable for over a month at ambient temperature, easily surviving the longest transit times from home to lab. GTR-STM enhances patient comfort, convenience, compliance and reduces infectious virus exposure to essential medical and lab professionals.

Alternatively, the GTR-STMdk direct-into-PCR product can be used to improve lab throughput and reduce reagent costs for saliva sample collection and testing at any lab site with access to refrigeration. GTR-STMdk reduces lab process time by 25% and reagent costs by 30% compared to other approaches. Since GTR-STMdk retains SARS-CoV-2 viral RNA stability for three days at ambient temperature, it is optimized for lab test site rather than at home saliva collection. SARS-COV-2 viral RNA levels as low as 0.4 genome equivalents/uL are detected in saliva samples using GTR-STMdk. The increased sensitivity of SARS-CoV-2 detection can expand COVID-19 testing to include asymptomatic individuals using pooled saliva.

**One Sentence Summary:** GTR-STM and Direct-into-PCR GTR-STMdk offer substantive improvements in SARS-CoV-2 viral RNA stability, safety, and RT-PCR process efficiency for COVID-19 testing by using a non-dilutive saliva sample collection system for individuals at home or onsite respectively.

## Introduction

The global COVID-19 pandemic, caused by the novel beta-coronavirus SARS-CoV-2, continues to ravage countries, with the United States bearing the brunt of the attack, and other nations bracing for a second wave (1-3). Testing and contact screening are the primary tools necessary to prevent and contain the infection within communities (4-8).

Rapid strides have been made in advancing molecular and antigen-based testing, which measure viral RNA and surface protein respectively (9-12) for the SARS-CoV-2. Significant progress has been achieved on three fronts, all of which play a significant role in improving patient comfort, patient convenience, and therefore, patient compliance. One major advancement is the shift away from complicated nasopharyngeal (NP) sampling to simpler saliva sampling.

CDC guidance recommends nasopharyngeal (NP) swabs for collection of samples for SARS-CoV-2 RT-PCR testing (13). NP sample collection can be uncomfortable, is not suitable for self-collection, and puts the health care professional at risk of infection (14,15). Some of the challenges for collection of NP sample include: (i) Difficulty in collection as the NP probe must be inserted deep into the nostril causing discomfort to the patient and often a gag/cough response. (ii) An element of risk for the trained healthcare professionals authorized to collect the NP sample (15,16). (iii) Shortages in availability of swabs and personal protective equipment for testing, a situation that led to significant under-testing of US populations at risk during the first few months of the pandemic (17,18). (iv) Requiring significant increases in the numbers of trained (currently insufficient) healthcare personnel to staff sample collection sites for testing to expand to mass screening.

In contrast to NP swabs, saliva is a convenient, safe, and effective sample for SARS-CoV-2 testing, even for children (19-22). This makes it possible to have home saliva collection kits in which patients self-collect the saliva in a sampling tube, which is then mailed to a CLIA-certified laboratory for testing and analysis. Recently, the FDA has approved saliva as an alternate sample for COVID-19 testing (23). Saliva yields diagnostic test results comparable to NP swabs (22). The ease of collection of saliva makes it ideal for the expansion of repeated testing programs in schools, workplaces, and for frequent travelers.

### Overview of Existing Saliva Sample Collection and Transport Media Products

Most saliva sampling and transport media products currently marketed require a two-step process for saliva collection. First, approximately 1mL of saliva must be expectorated (spat) into the sample tube. Second, an additional 1mL to 1.5mL of a sample-stabilizing liquid, supplied by the manufacturer, must be added to the saliva in the sampling tube (the rationale for this step is to preserve viral RNA integrity, and is discussed in detail in the next section).

The second step above; addition of sample-stabilizing liquid, adds problems that may adversely affect the quality of downstream laboratory results. (i) Reliance on an untrained user to add the secondary liquid introduces the potential for error. If missed, the quality of the sample and therefore the integrity of the test result may be compromised due to viral RNA degradation in transport. (ii) More problematic is the fact that the initial viral load in 1mL of collected saliva will be diluted by the additional stabilizing liquid, reducing the detectability of samples with low viral loads, increasing the potential to yield false negative results. (iii) The dilution problem will be particularly acute when mass screenings for SARS-CoV-2 are extended to asymptomatic individuals in schools, workplaces, and to travelers, many of whom may have low or very low levels of virus in saliva. Dilution of these samples by adding a secondary liquid can cause low positive samples to be missed. (iv) The issue of false negative results due to sample dilutive media will also be a severe problem with the adoption of pooled testing, where multiple samples may be combined with a low level positive in a pool, thereby impacting the Limit of Detection (LoD) of the assay.

### Assay Sensitivity and False Negative RT-PCR Test Results Due to viral RNA Degradation

Unlike a DNA sample (such as from a human genome test), which is very stable and maintains its molecular integrity under a variety of harsh conditions, RNA in saliva, which is RNAse-rich, is notoriously unstable and easily degraded. This accounts for the wide variation in test results reported in terms of high false negatives and LoD’s. To address this problem, assay protocols typically incorporate two additional steps before starting the PCR reaction. (i) Immediately after sample collection viral RNA stabilization media is added to ensure RNA integrity in the test samples. (ii) An aliquot of the diluted specimen is withdrawn, and the RNA is then subjected to an amplification process to ensure that sufficient quantities are available to serve as a PCR reaction template.

These additional sample treatment steps automatically bring with them more possibilities for error, potentially impacting the quality and accuracy of the test result. The ultimate solution is to find a way to eliminate these steps, by directly adding the undiluted and non-pre-treated saliva to the PCR reaction.

### Non-Dilutive Saliva Transport Media, With Less Pre-Analytical Processing Steps

Two devices that address the limitations of saliva sample collection described above are GTR-STM and GTR-STMdk. They were developed by GenTegra LLC, a Pleasanton, California-based company focused on developing products for ambient temperature stabilization and storage of RNA, DNA, proteins, and other biomolecules. The company’s Active Chemical Protection™ (ACP) technology incorporates a proprietary combination of small molecule inhibitors, cytotoxins, antioxidants, metal chelators, and anti-microbials to provide total protection for viral RNA at ambient temperatures. Proprietary ACP chemistry reduces saliva viscosity, inactivates contaminating RNases, and comprises ingredients that inactivate viruses and bacteria. The formulation renders the sample safe for transportation and testing. Furthermore, ACP is guanidinium-free, which makes it compatible with lab automation and self-sterilizing PCR instrumentation.

Both the GenTegra sample collection devices incorporate a non-dilutive dried media formulation at the bottom of the collection tube that eliminates sample dilution when oral fluid (saliva) is added by simply expectorating into the tube. The ionic and non-ionic denaturants in GTR-STM and the enzymes in GTR-STMdk inactivate the virus by disrupting the lipid envelope of the virus, rendering the saliva safe for transportation and testing. In order to further sterilize the saliva samples, the saliva in both GTR-STM and GTR-STMdk can be heated at 95°C for 15 minutes while still maintaining the integrity of the viral RNA for downstream RT-PCR analysis. Unlike EUA-approved home saliva kits available on the market, no additional liquid medium need be added to the saliva sample collected. The dried saliva transport medium in GTR-STM and GTR-STMdk dissolves upon contact with the saliva, thereby stabilizing the viral RNA. GTR-STM and GTR-STMdk makes a more concentrated and more stable RNA sample available for PCR, likely improving the assay LoD.

The distinct features and benefits of the two products are described below.

### GTR-STM™

- Optimized for home-based saliva sample collection
- For use with legacy high-throughput RT-PCR assay systems, which incorporate an RNA extraction and pre-amplification step
- Contains a proprietary anti-microbial RNA stabilization formula in dry form, thereby enabling non-dilutive collection of saliva samples eliminating the need for additional liquid media
- Comprises ingredients that inactivate virus in the sample by dissolution and denaturation rendering it safe for transport and testing
- Retains SARS-CoV-2 viral RNA stability with minimal RNA degradation for at least **60 days** at ambient temperature

### GTR-STMdk™

- Optimized for clinical site saliva sample collection that bypasses sample preparation step
- Contains Proteinase K in a proprietary formulation in dry form, enabling non-dilutive collection of saliva samples and eliminating the need for additional liquid media
- Comprises ingredients that inactivate live virus in the sample, rendering it safe for transport and testing.
- Retains SARS-CoV-2 viral RNA stability with minimal RNA degradation for up to **three days** at ambient temperature (the time typically required for saliva sample collection and Direct into PCR testing in a lab setting) and up to nine days at 4°C.
- Improves workflow due to:
  - Direct addition of saliva sample to RT-PCR process due to specially modified dry formulation
  - Eliminates need for sample pre-extraction step prior to RT-PCR assay, saving 25% in lab processing time
  - Can be used with sample volumes as low as 3.75uL and 30% less quantity of RT-PCR reagents
- Significant reduction in turnaround times for reporting assay results
- Significant reduction in testing cost by bypassing costs associated with sample extraction

The workflow using **GTR-STM** is summarized in **Figure 1, left side**. After saliva samples are received either from test-site or home collection, they are kept at ambient temperature until ready to process. They are then disinfected by heating at 95°C for 15 minutes in order to further protect lab personnel. The next step is a crucial prerequisite for successful RT-PCR. Aliquots (100 to 400 uL) of the sample are subjected to extraction using appropriate commercially available reagents (process time = nearly one hour). Then, 3.75 to 5uL of the extracted RNA sample is amplified in a total reaction volume of 10uL to 15uL of RT-PCR mix (process time = nearly one hour) followed by analysis. The entire lab test process including and up to recording of results in the system takes about four hours. (Note: the time from lab workflow process to actual delivery of assay results to patients varies from lab to lab and may take more than 24 hours after sample receipt.)

**Figure 1:**
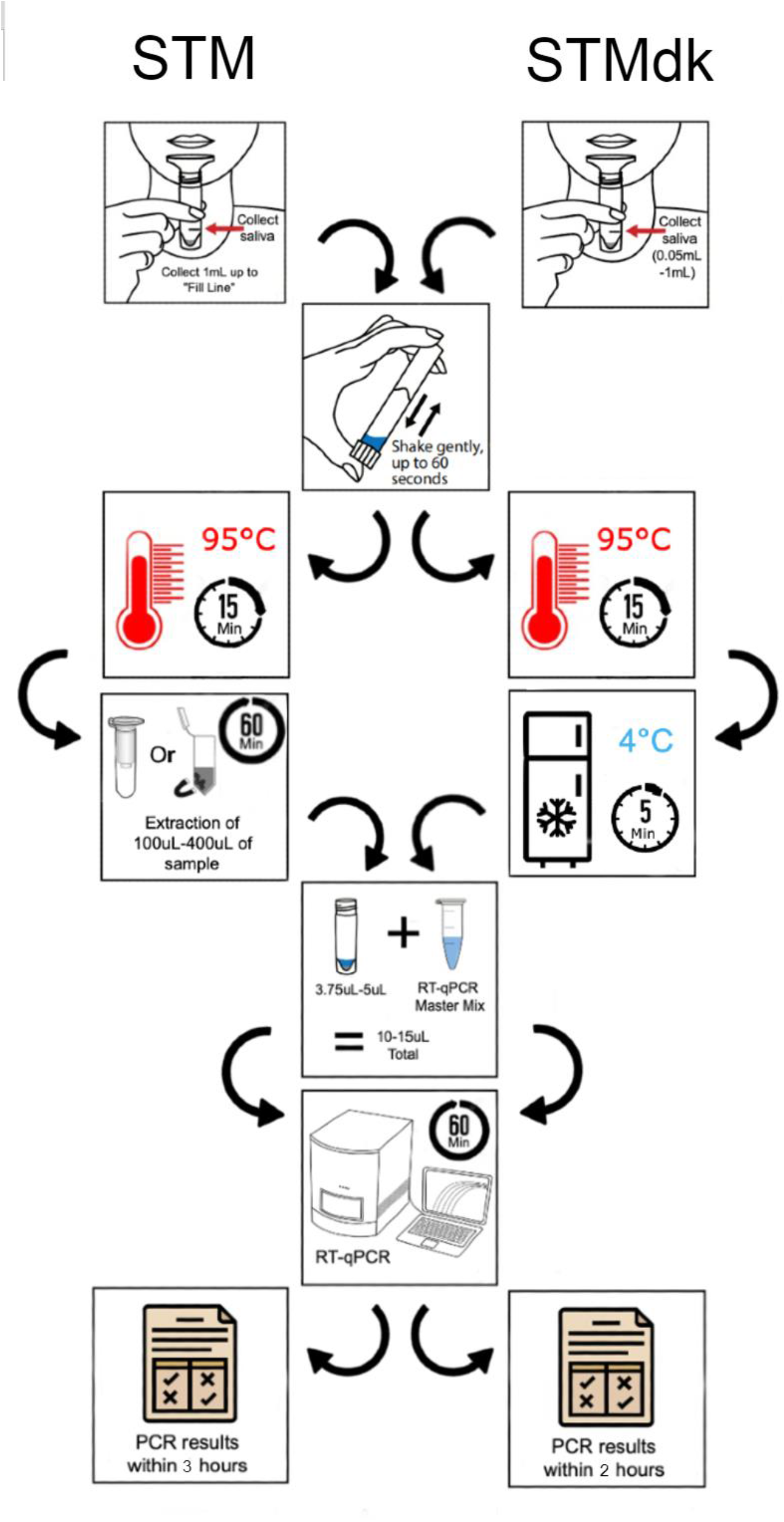
Workflow Comparison SARS CoV-2 RT-PCR Assay of Saliva Samples Collected in GenTegra GTR-STM vs. GTR-STMdk.

The workflow using **GTR-STMdk** is summarized in **Figure 1, right side**. Here, following saliva collection at the lab, the samples are kept at ambient temperature for 60 minutes to enable complete dissolution of the dried chemicals and activation of pre-dried Proteinase K (process time = nearly one hour). The samples are then heated for 15 minutes at 95°C (process time = nearly 15 minutes). This step will kill any residual Proteinase K that could potentially interfere with Taq polymerase during RT-PCR and simultaneously disinfect the samples to ensure lab personnel safety. The samples are then cooled at 4°C for five minutes before adding directly into the RT-PCR reaction system for assay and analysis (process time = nearly one hour). Since there is ***NO*** extraction and pre-amplification step, the entire process up to recording of results in the system takes about three hours.

## Results

CDC guidelines suggest using N1 and N2 primers for amplification in the RT-PCR assay for SARS-CoV-2. We ran a comparative analysis of N1 and N2 primer data using SARS-CoV-2-RNA spiked saliva and established by ANCOVA that the data obtained from either primer (N1 or N2) were statistically equivalent (Supplement S1). Therefore, due to cost and time considerations, all studies reported in this paper (except **Figure 6**) were performed using only the N1 primer from CDC.

### GenTegra-Saliva Transport Media (GTR-STM) - For Home Saliva/Sputum collection

Figure 2 compares RT-PCR data of gamma-irradiated SARS-CoV-2 virus spiked in saliva kept in GTR-STM device or non-GTR STM tube (“Saliva”). The data show comparable amounts of RNA are extracted (i.e., equivalent Cycle Threshold [**CT**] values were obtained) under either conditions.

**Figure 2:**
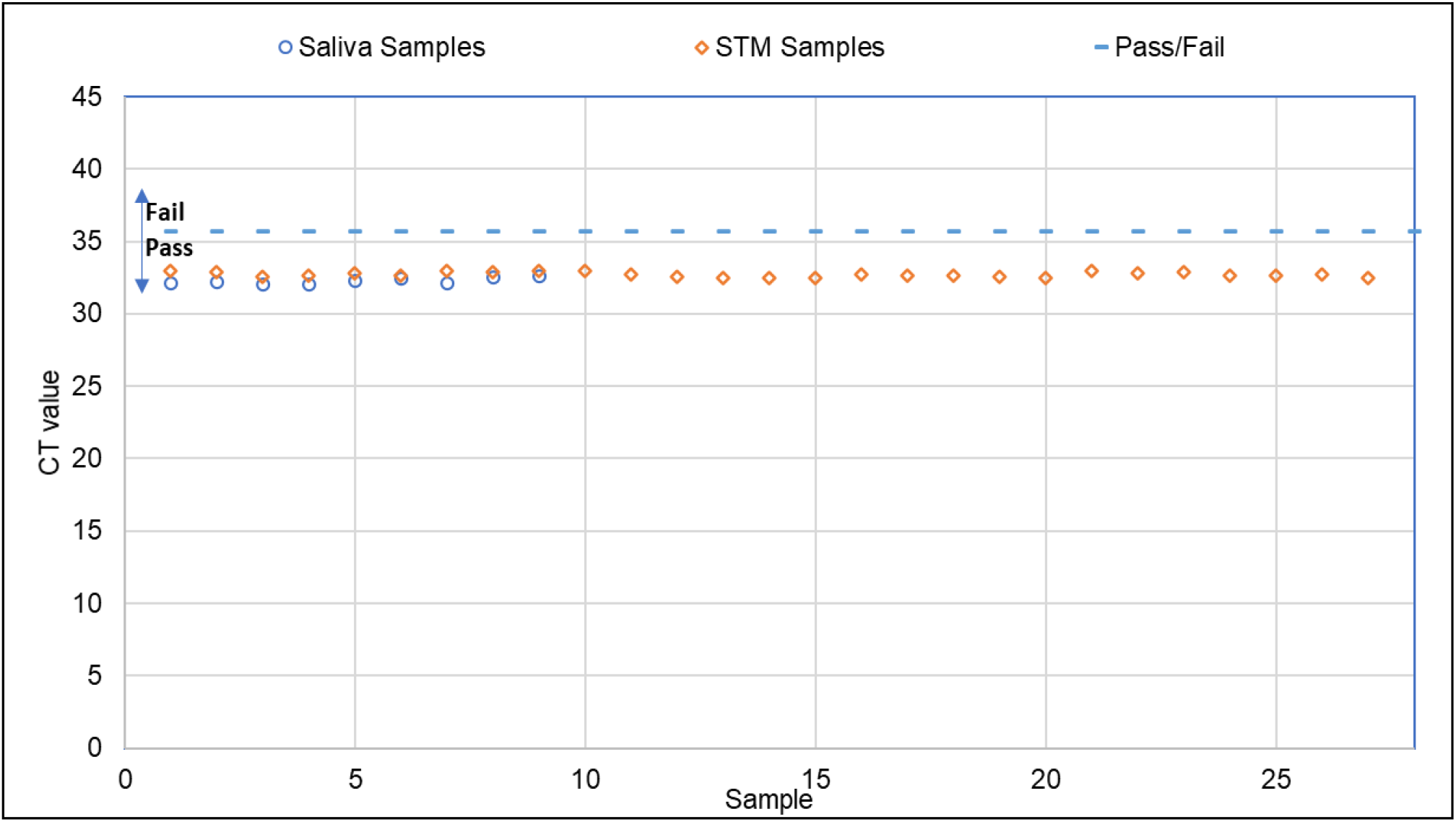
Extraction of Saliva-spiked gamma-irradiated SARS CoV-2 virus: “Saliva” v/s GTR-STM tubes. Gamma-irradiated SARS-CoV-2 virus (BEI Resources) at 2 genome equivalents/uL was spiked in 1mL of saliva kept either in non-GTR STM (“Saliva”) (open circles) or GTR-STM devices (open diamonds), extracted with MagMAX Viral RNA Kit (ThermoFisher) and RT-PCR performed with CDC’s N1 primer. The pass/fail criteria set at 35.7 CT is 3 CT values more than the average CT value of the “Saliva” only samples. “Saliva” samples without GTR-STM gave a mean CT of 32.4 CT (Std Dev, ±0.3), and Saliva Samples in GTR-STM gave a mean CT of 32.7 CT (Std Dev, ±0.2).

> **Study setup**
>
> <u>Experimental Sample</u>: A contrived GTR-STM sample (n=9) was prepared by spiking 1mL of saliva with 2 genome equivalents/uL of -gamma-irradiated SARS-CoV-2 virus.
>
> <u>Control sample</u>: A contrived non-GTR STM (“Saliva”) (n=3) sample was prepared by spiking 1mL of saliva with 2 genome equivalents/uL of gamma-irradiated SARS-CoV-2 virus.
>
> <u>Sample Extraction</u>:RNA was extracted from 200uL of sample from both control and stressed samples following manufacturer’s instructions for MagMAX Viral RNA (ThermoFisher) manual protocol and eluted with 50uL of elution buffer.
>
> <u>Quantification</u>: Amplify 5uL of extracted RNA from each sample in triplicates with TaqPath master mix (ThermoFisher) and CDC’s N1 Primer (IDT).

Saliva has a high concentration of RNase (24). When saliva samples containing SARS-CoV-2 are collected at home and shipped to the remote testing site, there is the likelihood that during transport at ambient temperatures, the viral RNA could be degraded due to exposure to raw saliva. However, the presence of proprietary ACP chemistry in GTR-STM protects and stabilizes the SARS-CoV-2 viral RNA (**Figure 3**).

**Figure 3:**
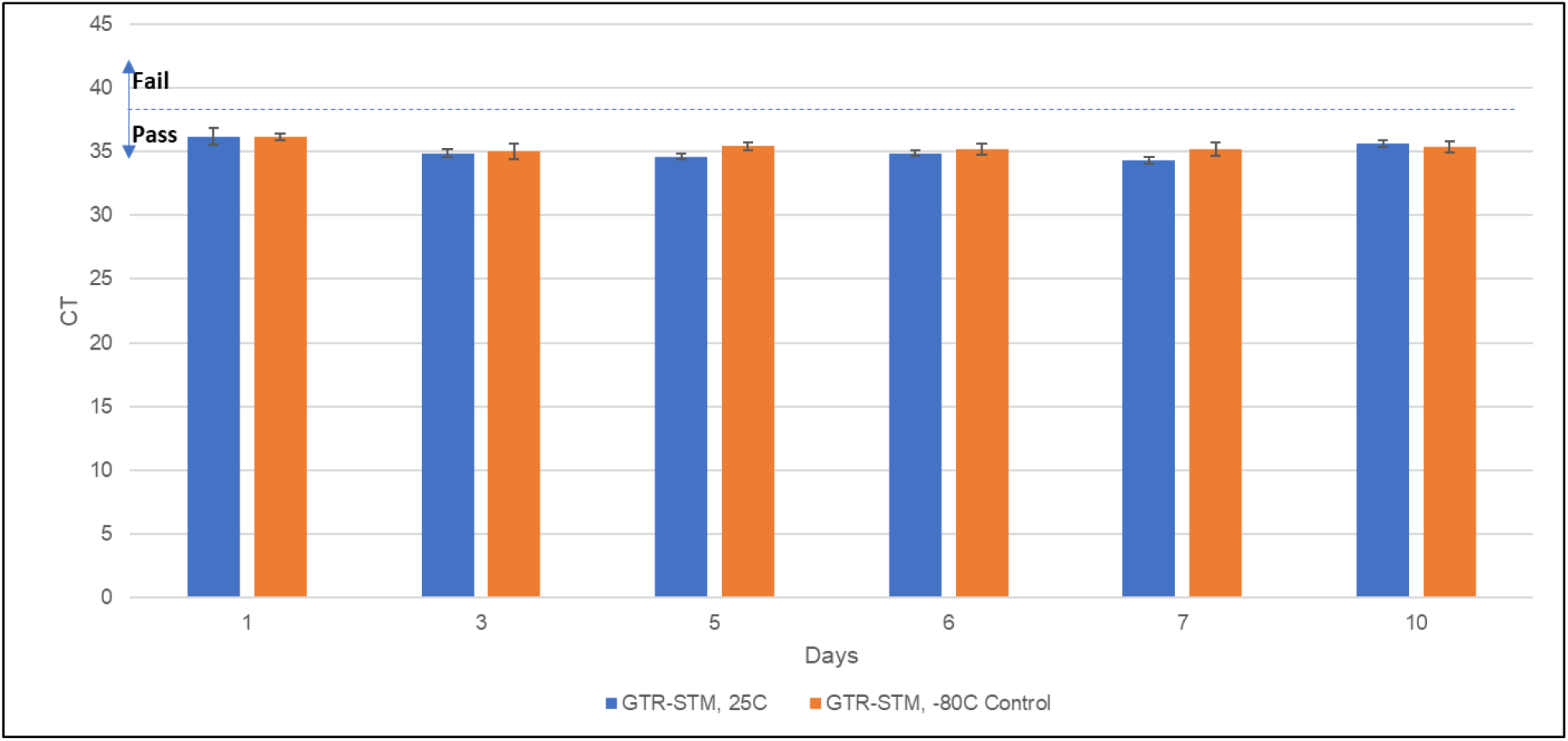
SARS-CoV-2 RNA (NOT whole virus)-spiked Saliva stabilized in GTR-STM™ for 10 days at ambient temperature (25°C) SARS-CoV-2 viral RNA (BEI Resources) at 3.0 genome equivalents/uL was spiked into 1mL of saliva kept in collection devices with and without GTR-STM, and stored at 25°C for 10 days. Matched spiked control saliva samples were stored at -80°C. No viral RNA was extracted from saliva without GTR-STM as the viral RNA is degraded immediately on spiking the saliva with SARS-CoV-2 RNA. Saliva in GTR-STM spiked with <u>viral RNA</u> and stored for up to ten days at 25°C, yielded ∼100% recovery (Figure 2, **blue bars**) compared to -80°C control (Figure 2, **maroon bars**). The pass/fail criteria for saliva in GTR-STM is 38.2 CT. Average CT for control (−80C) GTR-STM samples spiked with SARS-CoV-2 RNA is 35.2 ±0.4 and 36.6±0.3 for GTR-STM saliva samples spiked with SARS-CoV-2 RNA stressed at 25°C.

> **Study setup**
>
> <u>Experimental Sample</u>: 1 mL of Saliva sample added to dry GTR-STM + 3.0 geq/uL of SARS-CoV-2 viral RNA (BEI Resources). The viral RNA-containing saliva sample was incubated at 25*°*C for up to 10 days.
>
> <u>Control sample</u>: 1mL of RNAsecure water spiked with 3.0 geq/uL viral RNA and stored at -80°C.
>
> <u>Sample Extraction</u>: RNA was extracted from 100uL of experimental & control samples with QIAamp Viral RNA kit and eluted with 100uL of elution buffer.
>
> <u>Quantification</u>: 5uL of RNA was quantified with CDC’s SARS-CoV-2 RT-qPCR assay for N1 primer.

The stability studies were further extended using heat-inactivated whole SARS-CoV-2 virus (sourced from BEI resources) spiked into saliva in GTR-STM. When spiked into saliva with GTR-STM, the heat-inactivated SARS-CoV-2 virus was stable for up to 36 days at 25*°*C, and up to seven days at 56*°*C, prior to RNA extraction and RT-qPCR analysis (**Figures 4 and 5**).

**Figure 4:**
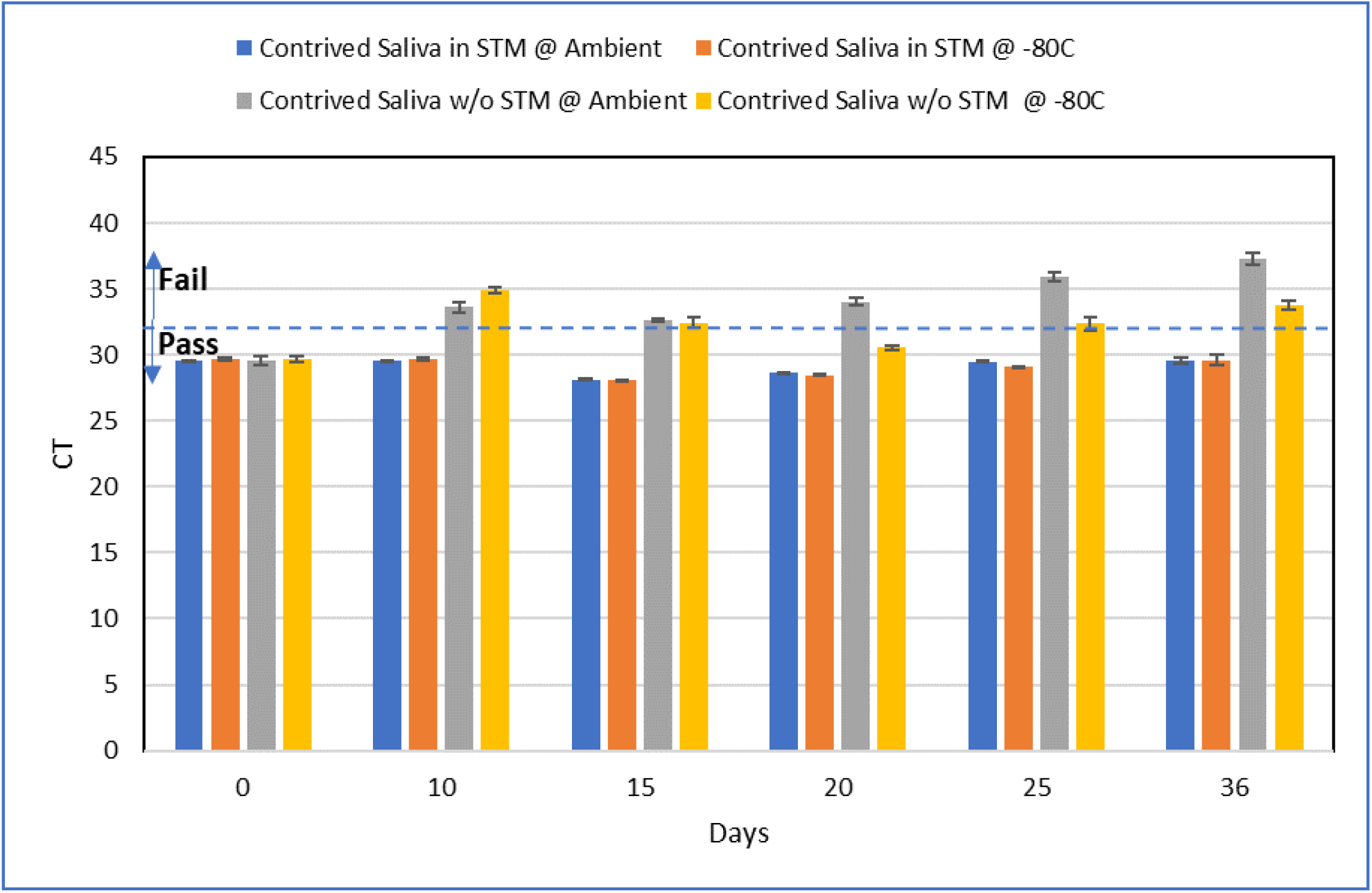
Stability of Saliva-spiked heat inactivated SARS CoV-2 virus stored in GTR-STM devices vs non-GTR-STM (“Saliva”) tubes at 25°C. Heat-inactivated SARS-CoV-2 virus RNA (BEI Resources) at 500 genome equivalents/uL was spiked into 1mL of saliva kept either in GTR-STM collection devices or non-GTR-STM (“Saliva”) tubes, and stored at 25°C for 36 days. Matched spiked control saliva samples in both kinds of tubes were stored at -80°C. The pass/fail criteria are set at 32 CT value. 200uL of the sample is used at each time point to extract viral RNA with the MagMAX kit with a final elution volume of 50uL. The CT value of the viral RNA extracted with MagMAX viral RNA kit is normalized to input volume of 200uL (volume of sample used for RNA extraction). All GTR-STM samples gave excellent recoveries when compared to their matched -80°C control. Both the -80°C and the 25°C samples for the non-GTR-STM (“Saliva”) is above the pass/fail line even in the -80°C control samples indicating that the viral RNA is degraded by the Rnase in the short time (less than half hour) that the saliva sample is defrosting before RNA extraction is performed.

> **Study setup**
>
> <u>Experimental Samples</u>: 1 mL aliquots of saliva contrived with SARS-CoV-2 at 500 genome equivalents/uL (BEI Resources) were placed into either GTR-STM devices or non-GTR-STM (“Saliva”) tubes and stored at ambient (25°C) for up to 36 days.
>
> <u>Control sample</u>: 1 mL aliquots of saliva contrived with SARS-CoV-2 at 500 genome equivalents/uL (BEI Resources) were placed into either GTR-STM devices or non-GTR-STM (“Saliva”) tubes and stored at -80°C for up to 36 days.
>
> <u>Sample Extraction</u>: RNA extracted from 200uL of experimental and control samples with MagMAX viral RNA kit at days 10, 15, 20, 25, and 36 and eluted in 50uL of elution buffer.
>
> <u>Quantification</u>: 5uL of RNA was quantified with CDC’s SARS-CoV-2 RT-qPCR assay for N1 primer.

**Figure 5:**
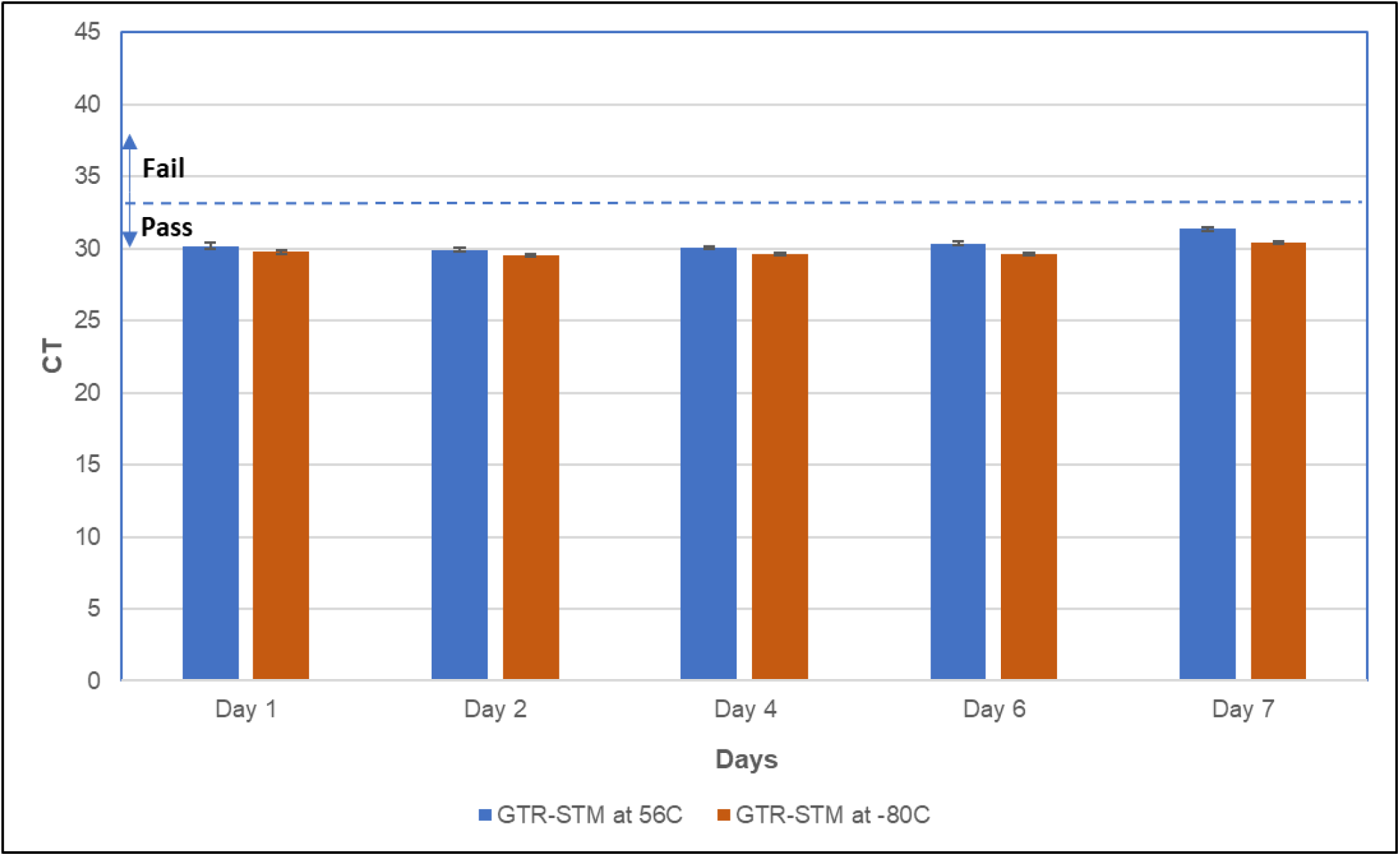
Stability of Saliva-spiked heat inactivated SARS CoV-2 virus in GTR-STM™ at 56°C. Heat inactivated SARS-CoV-2 viral RNA (BEI Resources) at 900 genome equivalents/uL was spiked into 1mL of saliva kept in collection devices with and without GTR-STM, and stored at 56°C for seven days. Matched spiked control saliva samples were stored at - 80°C. The pass/fail criteria is set at 32 CT values.

> **Study setup**
>
> <u>Experimental Sample</u>: 1 mL of saliva samples contrived with 900 geq/uL of heat inactivated SARS-CoV-2 Virus (BEI Resources) were stored in GTR-STM devices. The samples were incubated at 56°C for up to 7 days.
>
> <u>Control sample</u>: 1 mL of saliva samples contrived with 900 geq/uL of SARS-CoV-2 Virus (BEI Resources) were stored in GTR-STM devices at -80°C.
>
> <u>Sample Extraction</u>: RNA was extracted from 100uL of experimental and control samples with QIAamp Viral RNA kit and eluted in 100uL of elution buffer.
>
> <u>Quantification</u>: 5uL of RNA was quantified with CDC’s SARS-CoV-2 RT-qPCR assay for N1 primer.

SARS-CoV-2 virus samples stored in GTR-STM retained original viral RNA integrity (CT levels below the “Pass/Fail” level) for up to 36 days at ambient temperature, and the CTs values were comparable to controls stored at -80°C (**Figure 4**). In stark contrast, the samples stored in non-GTR-STM (“Saliva”) tubes did not meet the “Pass/Fail” CT level criterion at day 10 at ambient or on subsequent days. The ambient stability difference between GTR-STM and non-GTR-STM (“Saliva”) samples was shown to be statistically significant by ANCOVA (Supplemental Data, S2).

Surprisingly, under non-GTR-STM (“Saliva”) storage, even the -80°C stored samples showed inconsistent retention of CT levels below “Pass/Fail.” It is likely that in these frozen samples, SARS-CoV-2 viral RNA degradation occurred even during the very short five-minute thaw at 4°C prior to RT-PCR.

**Figure 5** shows an accelerated stability study by storing samples at an elevated temperature of 56°C.

Heat inactivated SARS-CoV-2 virus spiked saliva in GTR-STM retained its stability for up to seven days in GTR-STM. The data show indistinguishable CT values between samples stored at 56°C and -80°C, with all CT levels below the “Pass/Fail” criterion. This 56°C stability for seven days implies equivalent temperature stability at ambient (25°C) for stored samples of approximately 60 days, based on the Arrhenius equation for temperature dependence of reaction rates. Regression analysis performed (t-statistic test, P<0.05, R-squared value 0.625) based on the seven days data suggests that saliva containing SARS-CoV-2 virus collected in GTR-STM potentially be extrapolated to show stability at ambient temperature more than 214 days (Supplemental Data, S3).

The stability data in **Figures 4 and 5** demonstrate the utility of GTR-STM as a saliva sample collection device in a home setting, allowing for sufficient time from collection, shipping/transportation, and RT-PCR assay under highly variable temperature conditions.

Even though GTR-STM media contains components known to inactivate SARS-CoV-2, to ensure an added measure of safety of lab personnel, incoming sample specimens must be pre-heated at 95°C for 15-30 minutes to inactivate any residual live SARS-Cov-2 that may be present (25, 26). **Figure 6** shows that SARS-CoV-2 viral-spiked saliva samples in GTR-STM can be preheated at 95°C for up to 30 minutes prior to RT-PCR without adversely affecting assay sensitivity.

**Figure 6:**
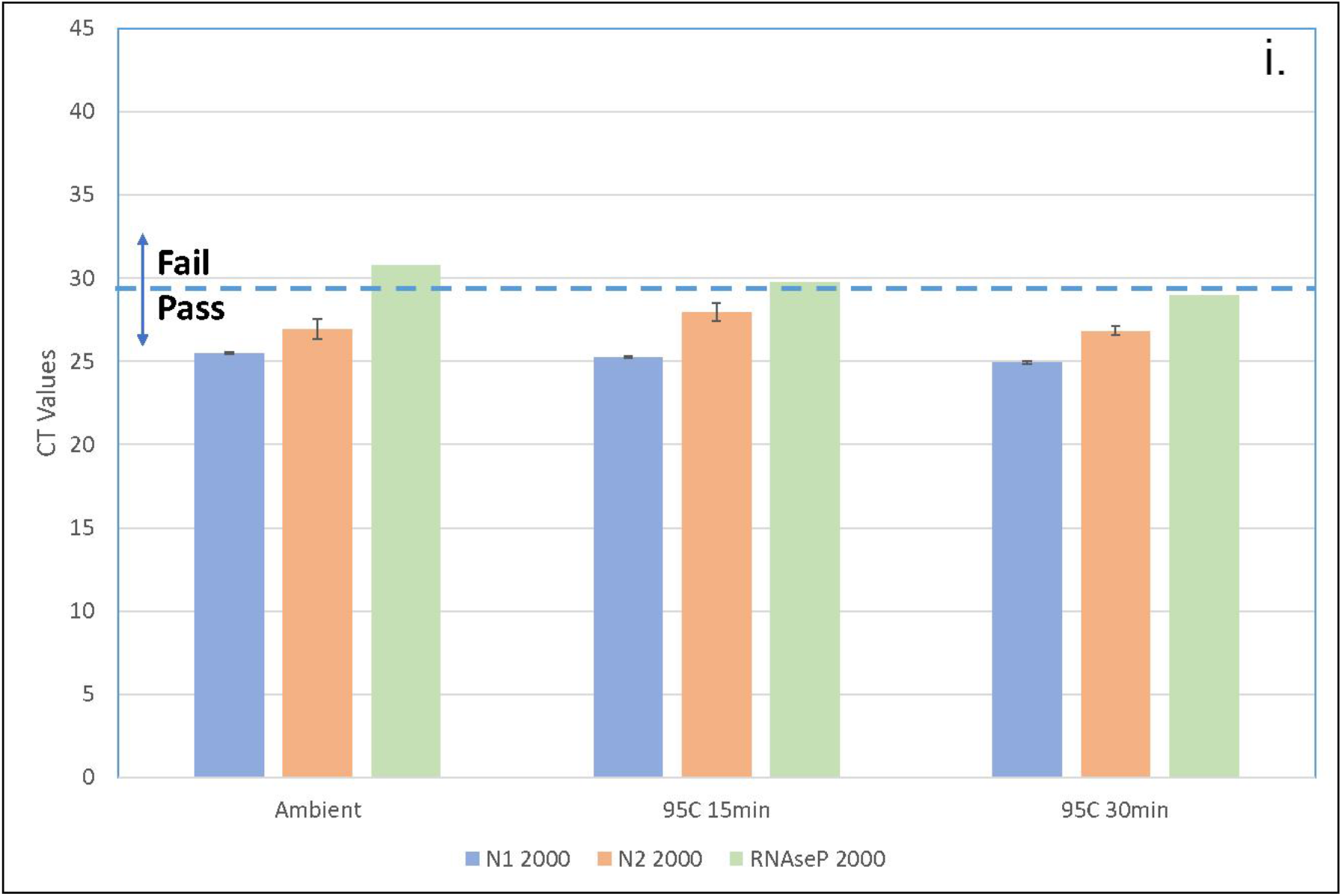
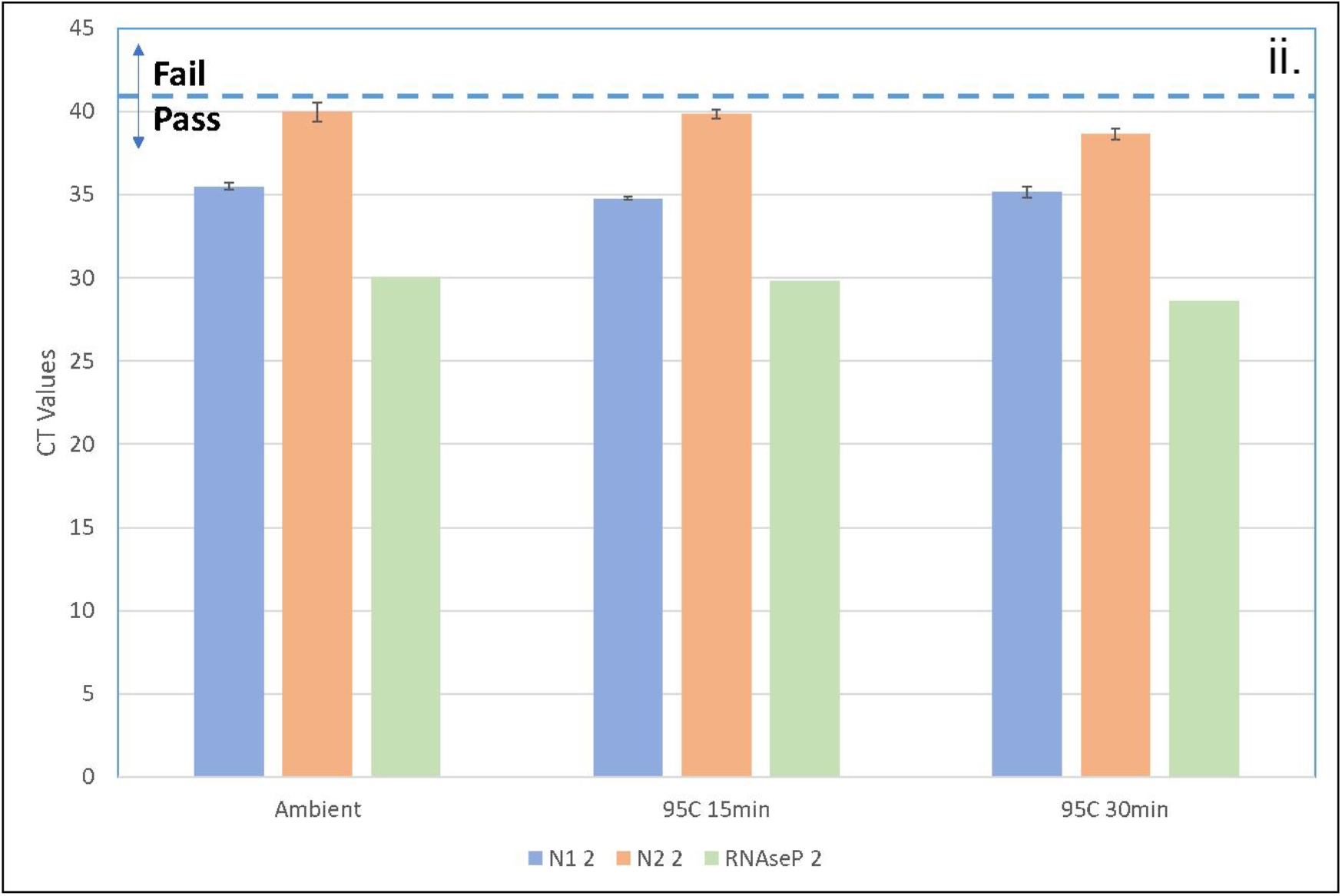
Effect of disinfection of gamma-irradiated SARS-CoV-2 spiked saliva samples collected in GTR-STM by preheating at 95°C. Gamma-irradiated SARS-CoV-2 virus (BEI Resources) at 2,000 (i.) and 2.0 (ii.) genome equivalents/uL was spiked in 1mL of saliva kept in GTR-STM, and heated at 95°C for up to 30 minutes. Matched spiked control saliva samples were kept at ambient temperature. The viral RNA was extracted with QIAamp Viral RNA Mini Kit and quantified with all three CDC primers, N1, N2 and RNase P. The pass/fail criteria of 29.22 CT values for 2,000 genome equivalent/uL and 40.74 CT values for 2.0 genome equivalents/uL were computed from the average of both N1 and N2 primers only at ambient temperature and not for RNaseP.

> **Study setup**
>
> <u>Experimental Samples</u>: Saliva sample was contrived with 2,000 and 2.0 geq/uL gamma-irradiated SARS-CoV-2 virus per uL. 1mL each of contrived samples were added to GTR-STM, heated at 95°C for up to 30 minutes.
>
> <u>Sample Extraction</u>: RNA was extracted from 50uL of sample with QIAamp viral RNA kit and eluted in 50uL of elution buffer.
>
> <u>Quantification</u>: 5uL of RNA was quantified by CDC’s N1, N2 and Rnase P primer sets.

The data in **Figure 6** showed that pre-heating of viral samples for 15 minutes or up to 30 minutes at 95°C gave CT values that were consistent with ambient samples using CDC’s N1, N2 or Rnase P primers for quantification. The N2 primer yielded higher CT values than N1. Perhaps the N1 primer binds to a more stable section of the SARS-CoV-2 viral RNA and therefore could be better suited to obtain greater sensitivity in RT-PCR testing for COVID-19.

All the above results (shown in **Figures 2 - 6**) clearly establish that GTR-STM is a valuable home saliva sample collection device since it (i) retains SARS-CoV-2 activity for 36 days at ambient temperature following sample collection and transportation to a testing lab, (ii) retains SARS-CoV-2 activity for up to seven days at 56°C, which translates to approximately 60 days at ambient temperature, (iii) can withstand heating conditions up to 56°C that may be encountered perhaps unexpectedly at transit points during shipping, and (iv) maintains viral RNA stability during disinfection by heating for up to 30 minutes at 95°C.

### GenTegra-Direct into PCR Media (GTR-STMdk) - For Lab Saliva/Sputum collection in Clinical Diagnostics Laboratory Settings

While the GTR-STM device solves the problem of dilution described earlier, diagnosticians are still left with the additional complication of the required extraction of viral RNA prior to PCR. Eliminating this step would significantly lower reagent costs per test, while also improving assay turnaround times, particularly when processing thousands of test samples. This second complication has also been resolved successfully with an improved device, GTR-STMdk.

Based on feedback from CLIA-lab partners, the “direct-into-PCR” product **GTR-STMdk**, eliminates the crucial 60 minute-pre-extraction and RNA amplification step. This is a significant advance as it saves up to 25% of workflow process time and reduces the added costs of non-PCR reagents. The key difference between GTR-STM and GTR-STMdk is the addition of Proteinase K pre-dried along with the RNA stabilization chemistry. Proteinase K degrades the viral coat, releasing the RNA and therefore eliminating the need of pre-extraction for RNA amplification.

The following experiments were performed with gamma-irradiated SARS-CoV-2 virus spiked saliva samples kept in GTR-STMdk, where the focus was to push the COVID-19 testing envelope in terms of improving process workflow and reagent cost efficiencies and increased sensitivity of SAR-CoV-2 detection. All experiments were conducted with clinically realistic low levels of SARS-CoV-2 spiked into saliva of 0.4 geq/uL and 2.0 geq/uL.

**Figure 7** demonstrates the effect of sample and reaction volumes on RT-PCR data using GTR-STMdk, an important consideration pointed out by CLIA lab partners. Since reagents account for one third of lab test costs, any reduction in total reaction volume used during the RT-PCR step, particularly when millions of samples are assayed, would yield substantial cost savings in testing.

**Figure 7:**
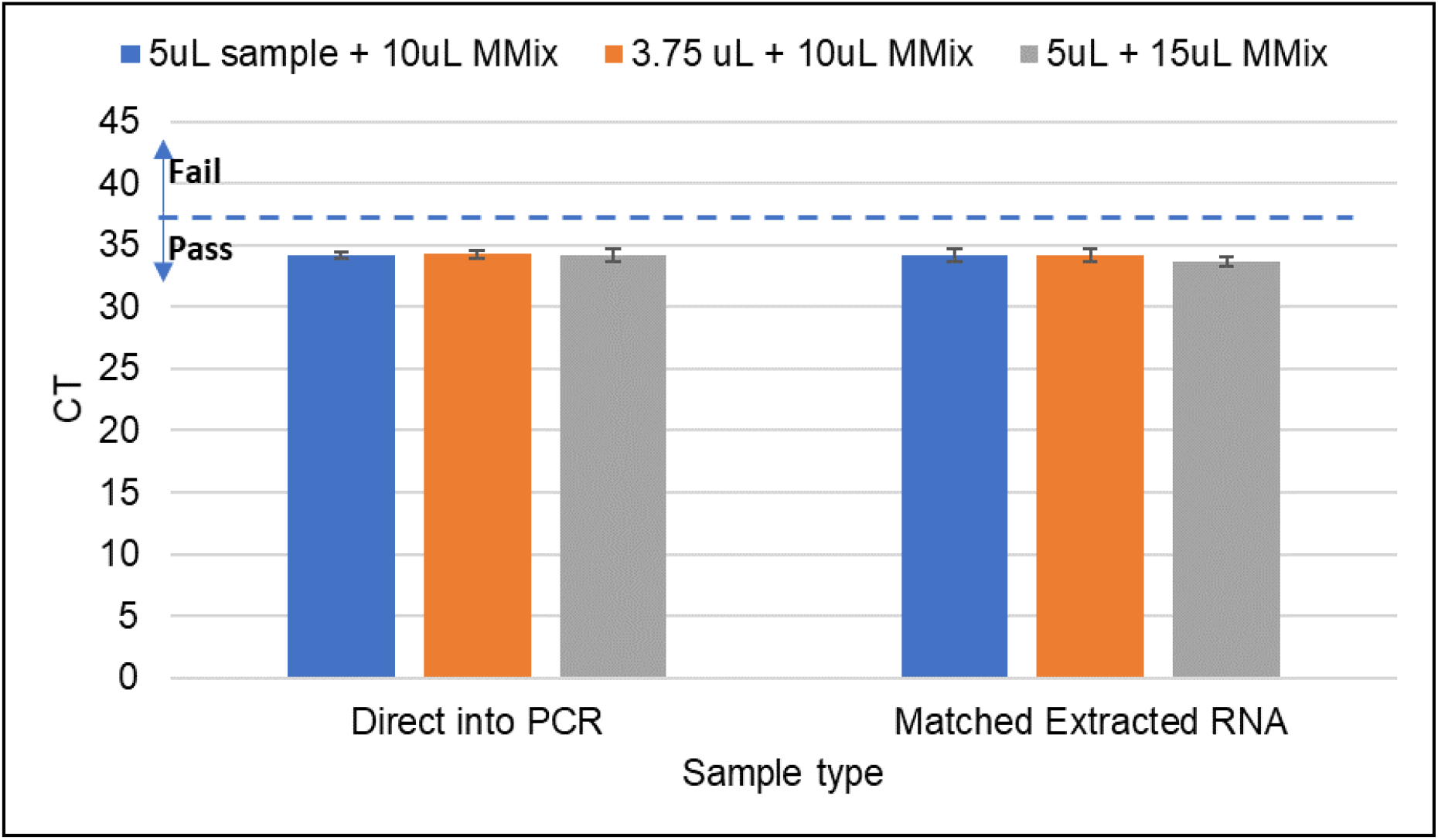
GTR-STMdk Sample Volume Titration in RT-PCR. 500uL of saliva spiked with 2.0 geq/uL of gamma-irradiated SARS-CoV-2 virus added to GTR-STMdk was incubated at 56°C for 5 minutes followed by 95°C for 5 minutes. Direct into PCR was performed with 3.75uL to 5.0uL of GTR-STMdk sample added to 10uL to 15uL of mastermix and amplified with CDC’s N1 primer. The pass/fail criteria set to Matched Extracted RNA samples at 37.2 CT.

> **Study setup**
>
> 500uL of saliva contrived with gamma-irradiated SARS-CoV-2 virus, 2 geq /uL, was added to GTR-STMdk device. Six replicates each with; (i) 5uL of sample to 10uL of TaqPath RT-PCR master mix, (ii) 3.75uL of sample to 10uL of master mix, (iii) 5uL of sample to 15uL of master mix (ThermoFisher volume).
>
> <u>Experimental Samples</u> were added directly into the RT-PCR assay after first heating for five minutes at 56°C, followed by 15 minutes at 95°C, and then cooling for 5 minutes at 4°C (“**Direct into PCR**” in Figure 8).
>
> <u>Control Samples</u> were extracted with QIAamp viral RNA kit from 100uL of saliva sample. The extracted RNA was eluted in 100uL of Elution Buffer (CT, STMdk Extracted), then subjected to RT-PCR (‘**Matched Extracted RNA**” in Figure 7)
>
> <u>Quantification</u>: 3.75uL to 5uL of RNA was quantified with CDC’s SARS-CoV-2 RT-qPCR assay for N1 primer.

The results show that similar CT values were obtained using a total RT-PCR volume of 13.75uL comprising 3.75uL test sample and 10uL of reagent master mix versus 20uL comprising 5uL test sample versus 15uL reagent master mix. Thus, potential savings in reagent costs of up to 30% could be realized using GTR-STMdk.

The practical utility of the GTR-STMdk device was further explored to account for variable volumes of saliva samples collected on site. COVID-19 patients may experience difficulties in generating sufficient saliva volumes due to their weakened condition. **Figure 8** shows RT-PCR data in which contrived saliva sample volumes of 50uL, 100uL, 500uL and 1,000uL were placed inside GTR-STMdk devices.

**Figure 8:**
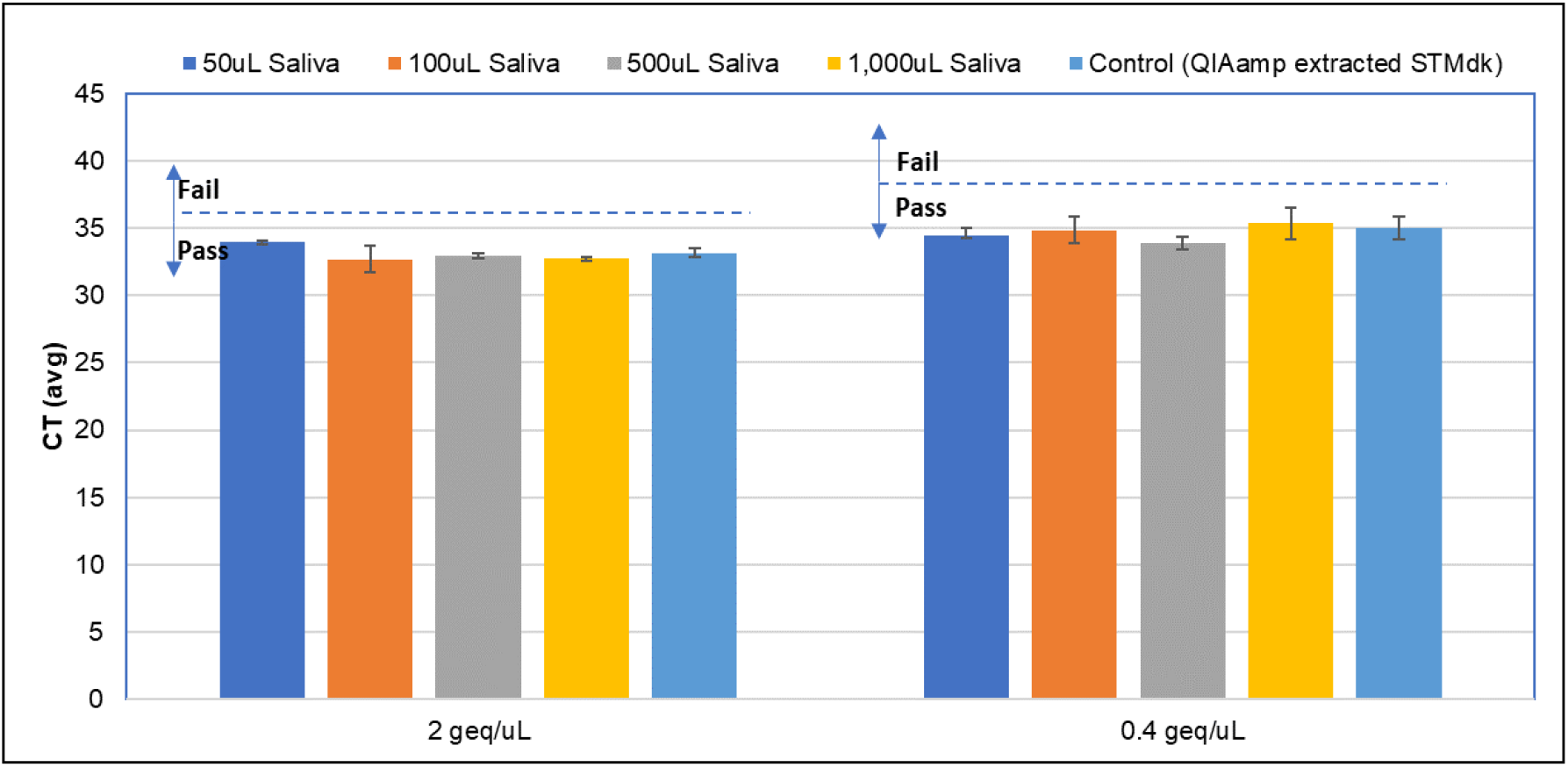
Saliva Volume Titration in GTR-STMdk. Various volumes (50uL to 1 mL) of saliva, spiked with gamma-irradiated SARS-CoV-2 virus (BEI Resources) at 0.4 genome equivalents/uL (Right Panel) or 2.0 genome equivalents/uL (Left Panel), were kept in GTR-STMdk before direct RT-PCR assay. The pass/fail criteria are set to the QIAamp extracted STMdk control samples for 2.0 geq/uL panel and 0.4geq/uL and is 36 and 38 CT values respectively.

> **Study setup**
>
> Various volumes, 50uL, 100uL, 500uL and 1,000uL of Contrived Saliva at 0.4 geq/uL and 2.0 geq/uL of gamma-irradiated SARS-CoV-2 (BEI Resources) added to STMdk devices.
>
> <u>Experimental Samples</u> were heated for 5 minutes at 56°C, then heated for 5 minutes at 95°C, cooled for 5 minutes at 4°C, then subjected to direct RT-PCR.
>
> <u>Control Samples</u> were extracted with QIAamp viral RNA kit from 100uL of saliva sample. The extracted RNA was eluted in 100uL of Elution Buffer (CT, STMdk Extracted).
>
> <u>Quantification</u>: 5uL of RNA was quantified with CDC’s SARS-CoV-2 RT-qPCR assay for N1 primer.

At both the low and high doses of SARS-CoV-2, samples at all saliva volumes tested gave CT values below the “Pass/Fail” criterion. Therefore, the GTR-STMdk device would be suitable for use with saliva sample volumes as low as 50uL and as high as 1mL.

Also, compared to the samples in **Figure 7**, the direct to PCR samples in **Figure 8** were heated for 15 minutes at 95°C without first being heated at 56°C for five minutes. The elimination of the 56°C step had no impact on CT “Pass/Fail” results at both SARS-CoV-2 virus levels - 0.4 geq/uL and 2.0 geq/uL.

As shown in the process workflow chart in the Introduction in **Figure 1**, in a typical clinical lab situation, so long as the saliva samples in GTR-STMdk are kept at ambient temperature for one hour before processing, only the 95°C heating step would be required. The 56°C step may be needed in a point of care diagnostic situation when saliva is collected and processed immediately.

**Figure 9** shows RT-PCR data of saliva samples spiked with gamma-irradiated SARS-CoV-2 at 0.4 geq/uL (Panel A) and 2 geq/uL (Panel B) in the direct-to-PCR GTR-STMdk device, with ***NO*** pre-extraction or -amplification. The samples were further assayed on days 1, 2 and 3 following storage at ambient temperature.

**Figure 9:**
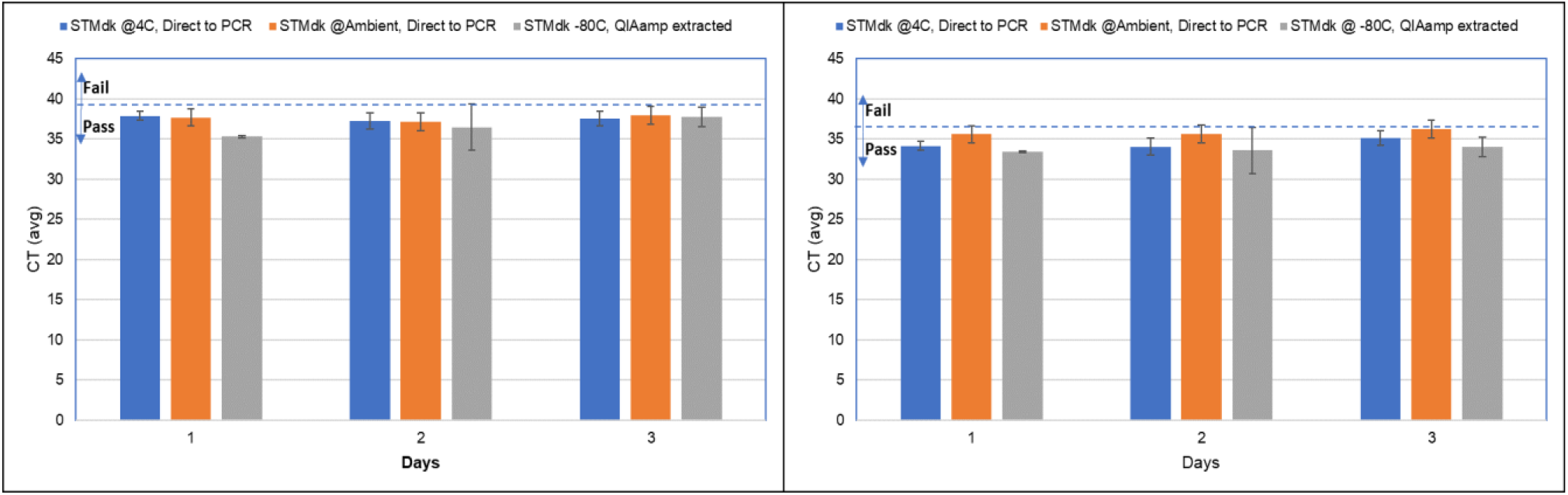
Stability of Saliva-spiked gamma-irradiated SARS CoV-2 virus in GTR-STMdk™ at 25°C. Gamma-irradiated SARS-CoV-2 virus (BEI Resources) at 0.4 genome equivalents/uL (Left Pane A) or 2.0 genome equivalents/uL (Right Panel B) was spiked in 500uL of saliva kept in GTR-STMdk, and stored at 4°C, 25°C, or -80°C for up to three days before direct RT-PCR assay.

> **Study setup**
>
> <u>Experimental Sample</u>: 500uL of Contrived Saliva at 0.4 geq/uL and 2.0 geq/uLof gamma-irradiated SARS-CoV-2 (BEI Resources) added to STMdk devices. They were stored at 4°C and 25°C.
>
> <u>Control Sample</u>: Two controls were set up in this experiment. One for direct into PCR was placed at 4°C in order to keep the Proteinase K active under ideal conditions and another set of controls for determining the amount of viral RNA that is to be expected for the direct into PCR sample. The later was determined by extraction of the viral RNA from this sample with QIAamp viral RNA kit (true control). Controls are 500uL of Contrived Saliva at 0.4 geq/uL and 2.0 geq/uL of SARS-CoV-2 (BEI Resources) added to STMdk devices. QIAamp extraction of viral RNA was from 100uL of -80°C control saliva samples and eluted with 100uL of elution buffer.
>
> <u>Sample Treatment</u>: 50uL aliquots from each experimental and control sample were withdrawn on days 1, 2 and 3, heated for 15 minutes at 95°C, cooled for 5 minutes at 4°C, then subjected to RT-PCR
>
> <u>Quantification</u>: 5uL of RNA was quantified with CDC’s SARS-CoV-2 RT-qPCR assay for N1 primer.

At both the low and high doses of SARS-CoV-2, all samples gave CT values below the “Pass/Fail” criterion up to three days. Saliva samples contrived with heat inactivated or gamma-irradiated virus kept in GTR-STMdk beyond three days at 25°C had CT values above the “Pass/Fail” threshold.

Independent studies by an external collaborator with SARS-CoV-2 infected clinical samples from COVID-19 patients collected in GTR-STMdk have demonstrated stability of the SARS-CoV-2 viral RNA for up to nine days at 4°C (data not shown). Therefore, the GTR-STMdk device is ideally suited for sample collection at testing sites, such as clinical labs, where the device can be quickly refrigerated following saliva sample collection.

## Discussion and Concluding Remarks

Many countries, including US, are still in the grips of the COVID-19 pandemic while other countries are experiencing a resurgence or a potential second wave. Mass screening for the presence of SARS-CoV-2 in the general population has become an imperative. GTR-STM and GTR-STMdk provide essential technological advances in COVID-19 testing, by enhancing the ease of sample collection, increasing assay sensitivity, improving safety and facilitating lab workflow while enabling lower costs and faster turnaround times.

Saliva sampling is a safer, more convenient and practical alternative to nasopharyngeal swabs. GenTegra has been on the forefront of developing dried-media sample collection devices for a variety of testing applications, and this technology has been logically extended to develop two non-dilutive saliva collection products, GenTegra **GTR-STM** and **GTR-STMdk**. GenTegra’s proprietary Active Chemical Protection chemistry is particularly applicable to SARs-CoV-2 PCR assays where prevention of RNA degradation and preservation of stability during transport from sample collection to lab test sites is essential to ensuring high test accuracy and sensitivity.

**GTR-STM**, while a simple device, has the following attributes: (i) direct collection of saliva by expectoration into a tube containing dried stabilizers to avoid sample dilution, (ii) use of stabilizers that are generally known to inactivate infectious virus, and (iii) SARS-CoV-2 stability for up to 60 days at ambient temperature. At the test site, the viral RNA in the sample would need to be extracted and amplified before PCR analysis. Due to its ability to retain viral RNA stability for several weeks at ambient temperature, **GTR-STM would be ideally suited for home-based sample collection after which the sample containing devices may be shipped and transported worldwide at temperatures up to 56°C to authorized CLIA labs**..

**GTR-STMdk** incorporates Proteinase K into the saliva sample collection device itself, eliminating the Proteinase K addition step at the testing site, which reduces potential avenues for error. In addition, the sample can be directly subjected to RT-PCR <u>without</u> the need for RNA pre-amplification. The benefit of **NOT** requiring RNA extraction and amplification at the test site provides a substantial improvement to throughput by reducing test turnaround times. In SARS-CoV-2 spiked saliva samples collected in GTR-STMdk, virus levels down to 0.4 geq/uL can be detected using as little as 3.75uL of sample volume and 30% less quantity of expensive PCR assay reagents. The device is also effective with saliva collection volumes as low as 0.05 mL, minimizing difficulty for patients struggling to provide a usable sample when under stress. **GTR-STMdk is ideally suited for sample collection at locations proximal to the testing site** as it requires sample storage at 4°C. Storage of GTR-STMdk saliva samples at the testing site at 4°C extends stability up to nine days (confidential personal communication from a CLIA partner, data not shown).

A comparison of the characteristics and utility of the two devices is given in **Table 1**.

**Table 1:**
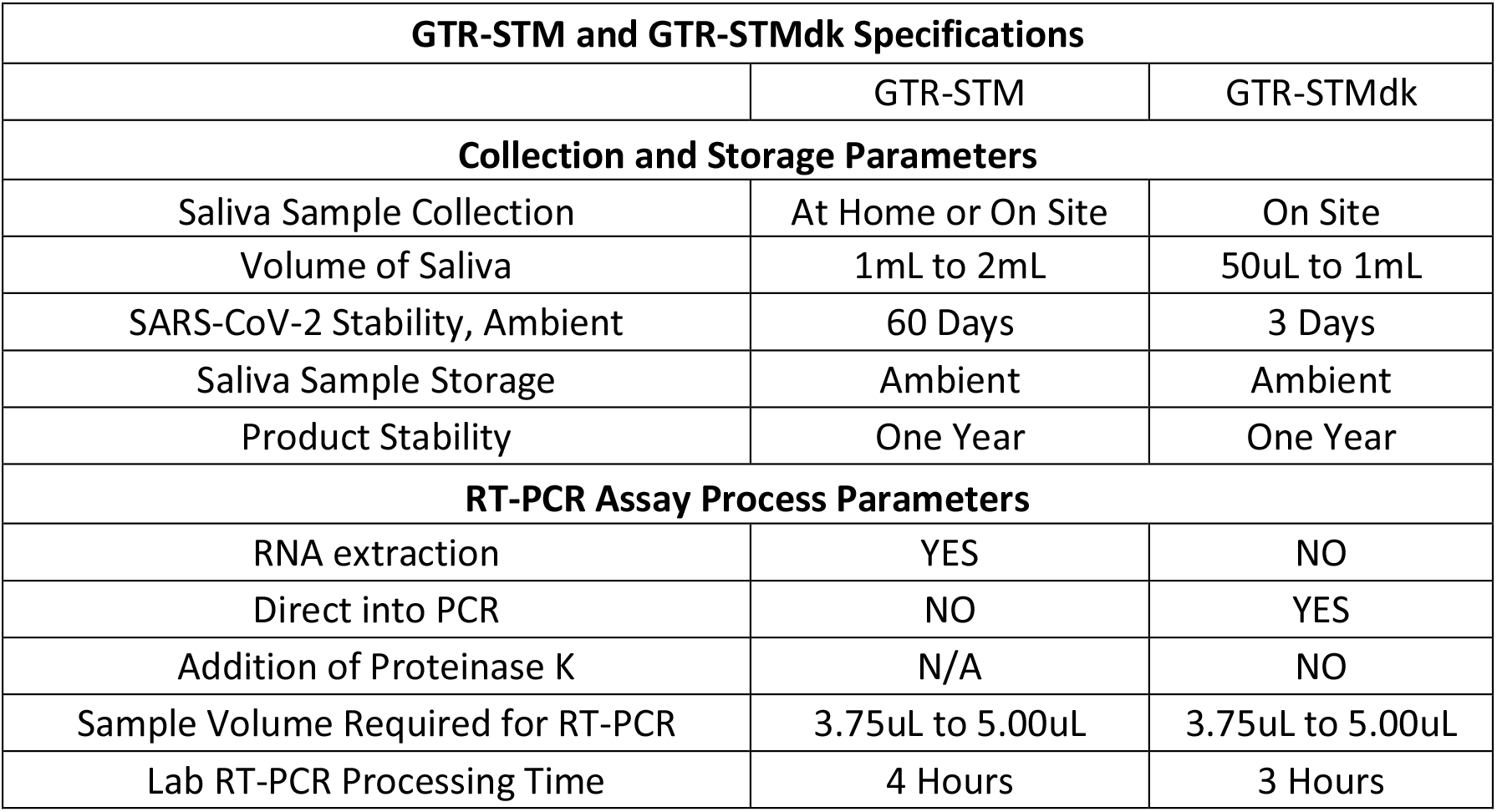
GTR-STM and GTR-STMdk Specifications.

| **GTR-STM and GTR-STMdk Specifications** |  |  |
| --- | --- | --- |
|  | GTR-STM | GTR-STMdk |
| **Collection and Storage Parameters** |  |  |
| Saliva Sample Collection | At Home or On Site | On Site |
| Volume of Saliva | 1mL to 2mL | 50uL to 1mL |
| SARS-CoV-2 Stability, Ambient | 60 Days | 3 Days |
| Saliva Sample Storage | Ambient | Ambient |
| Product Stability | One Year | One Year |
| **RT-PCR Assay Process Parameters** |  |  |
| RNA extraction | YES | NO |
| Direct into PCR | NO | YES |
| Addition of Proteinase K | N/A | NO |
| Sample Volume Required for RT-PCR | 3.75uL to 5.00uL | 3.75uL to 5.00uL |
| Lab RT-PCR Processing Time | 4 Hours | 3 Hours |

**Table 2** provides a comparison of the attributes of GTR-STM and GTR-STMdk to other saliva collection products currently marketed.

**Table 2:** GTR-STM and GTR-STMdk comparative saliva collection devices.

| Company | Gen Tegra | Gen Tegra | NORGEN | SPECTRUM | DNAGenotek | ZYMO | YALE UNIVERSITY | ISOHELIZ |
| --- | --- | --- | --- | --- | --- | --- | --- | --- |
| Product | GenTegra GTR-STM* | GenTegra GTR-STMdk* | Saliva RNA Collection and Preservation Device | SDNA 1000 | OMNIGene Oral Saliva, RNA & DNA collection | Shield™ saliva collection kits | SalivaDirect | GeneFIX™ Saliva DNA/RNA Collection |
| Requires Addition of Stabilizing Liquid Media? | NO | NO | YES | YES | YES | YES | YES | YES |
| Non-Dilutive? | YES | YES | NO | NO | NO | NO | NO | NO |
| Requires RNA Pre-Amplification Before RT-PCR? | YES | NO | YES | YES | YES | NO | NO | YES |
| Improves Workflow Efficiency? | NO | YES | NO | NO | NO | NO | YES | NO |
| Decreases Reagent Costs? | NO | YES | NO | NO | NO | NO | YES | NO |
| Sample Stability | • Ambient, 36 DAYS<br>• 56°C, 7 DAYS<br>(Equivalent to Ambient, 60 DAYS) | • Ambient, 3 DAYS | • Ambient, 28 DAYS | • Ambient, 14 DAYS | • Ambient, 21 DAYS | • Ambient, 28 DAYS | • Not Available | • Ambient, 14 DAYS |
| Product Shelf-life | • 24 months | • 24 months | • 24 months | • 30 months | • NA | • NA | • Not Available | • NA |
| * Please Note: GTR-STM and GTR-STMdk are available to CLIA certified laboratories and IVD assay kit manufacturers, pursuant to regulatory approvals in their respective US FDA Emergency User Authorization (EUA) protocols. |  |  |  |  |  |  |  |  |

In this report, the saliva used was from a commercially available source. Studies are in progress in GenTegra labs as well in collaboration with CLIA lab partners to confirm these data using individual human saliva samples. We have provided a conservative estimate for assay sensitivity, pending a more formal determination of test Limit of Detection (LoD) for samples collected in GTR-STM and GTR-STMdk.

The switch to dry chemical saliva sample collection devices GTR-STM and GTR-STMdk, can achieve significant improvements in SARS-CoV-2 RT-PCR testing throughput efficiencies. Saliva sampling with GTR devices also greatly enhances patient convenience, safety, reduces testing costs and reporting times for test results. The enhancement to accuracy, sensitivity and PCR assay turnaround times using GTR devices can greatly expand the number of asymptomatic individuals that can be tested on a regular basis while facilitating testing on pooled samples. The numerous sample testing advances provided by GTR devices are essential to support COVID-19 testing for all, with the ultimate critical goal to better control the ongoing pandemic until a safe and efficacious viral vaccine and therapies becomes widely available.

## Data Availability

Data is available on data.mendeley.com. Both GTR-STM and GTR-STMdk data and supplemental data has been uploaded to data.mendeley.com.

http://dx.doi.org/10.17632/tf57gxmxpd.1

## Abbreviations

COVID-19: Coronavirus Disease 19
SARS-CoV-2: Severe Acute Respiratory Syndrome Coronavirus 2
RT-PCR: Reverse Transcriptase-Polymerase Chain Reaction
PCR: Polymerase Chain Reaction
ANCOVA: Analysis of covariance
GTR-VTM: GenTegra Viral Transport Medium
GTR-STM: GenTegra Saliva Transport Medium
GTR-STMdk: GenTegra Saliva Transport Medium Direct Proteinase K
ACP: Active Chemical Protection
CLIA: Clinical Laboratory Improvements Act (1984)
RNA: Ribonucleic Acid
uL: Microliter
mL: Milliliter
geq/uL: Genome Equivalents per microliter
LoD: Limit of Detection
NP: Nasopharyngeal
CT: Cycle Threshold
CDC: Centers for Disease Control, Atlanta, GA
Rnase: Ribonuclease
MMix: Master mix

## Materials and Methods

Study setup for each set of experiments is given in the respective legend below each Figure.

Neat saliva samples were prepared by contriving commercially purchased saliva, (Lee BioSolutions, cat #991-05-P-250) with SARS-CoV-2 virus to the desired concentration. Heat-inactivated SARS-CoV-2 virus (BEI Resources, cat #NR-52286) at 1.12e^6 geq/uL and gamma-irradiated virus (BEI Resources, cat #NR-52287) at 1.7e^6 geq/uL. Unless otherwise stated in the respective Figure, a total of 1000 uL of contrived saliva was added to GTR-STM tubes and 500 uL of contrived saliva was added to GTR-STMdk tubes. GTR-STM tubes were gently mixed at room temperature for 5 minutes or until the pellet had gone completely into solution. GTR-STMdk tubes were allowed sit for 1 hour at room temperature.

GTR-STM was tested by extraction with either QIAamp Viral RNA Mini Kit (Qiagen, cat #52906) or MagMAX Viral RNA Isolation Kit (ThermoFisher, cat #AM1939) and compared with either PBS or neat saliva as controls. Samples extracted with QIAamp Viral RNA Mini Kit always contained either 50 or 100 uL of sample volume and were normalized by eluting in either 50 or 100 uL of elution buffer, depending on the sample volume. Samples extracted with MagMAX Viral RNA Isolation Kit followed a modified protocol and always contained 200 uL of sample volume and were eluted in 50 uL of elution buffer. The eluted viral RNA was then quantified with RT-qPCR (see below).

Two different heating methods were tested with contrived saliva in GTR-STMdk. First, samples were heated at 56°C for 5 minutes, before being heated at 95°C for 5 minutes. The second method tested was heating at 95°C for 15 minutes to deactivate the Proteinase K in the solution. The second method was sufficient in deactivating the Proteinase K before RT-qPCR and provided similar results to the first method, so the 56°C for 5 minutes step was removed.

Viral RNA samples from contrived saliva in GTR-STM and contrived saliva in GTR-STMdk were both quantified with RT-qPCR using a StepOnePlus™ Real Time PCR System (ThermoFisher Scientific). Samples were prepared, according to the CDC 2019-Novel Coronavirus (2019-nCoV) Real-Time RT-PCR Diagnostic Panel, by mixing 5 uL of viral RNA with 3.5 or 5 uL of TaqPath™ 1-Step RT-qPCR Master Mix, CG (ThermoFisher, cat #A15299), uL of HyPure™ Molecular Biology Grade Water (HyClone™, cat #SH30538.03), and 1.5 uL of CDC combined primer/probe mix (Integrated DNA Technologies, cat #10006713). CDC primer/probe mix includes N1, N2, and RNASEP. Samples were plated in triplicates into a 96-well plate. Thermocycler conditions consisted of 2 minutes at 25°C, 15 minutes at 50°C, 95°C at 2 minutes, and 45 cycles of 95°C at 3 seconds and 55°C at 30 seconds.

### Criteria for determination of SARS-CoV-2 positivity in RT-PCR Assay

Based on FDA guidance, samples were considered positive if the CT value was less than the “Pass/Fail” criterion of Mean CT + 3CT, and negative if above the “Pass/Fail” criterion.

## Acknowledgement

The authors would like to thank Dr. Raymond Lenhoff PhD. (Molecular Virologist, consultant), and Dr. Michael Hogan PhD. (CSO, PathogenDx), and Robert Barrette (VP Business Development, GenTegra LLC) for their insightful comments and edits.

